# A Rapid and Low-Cost protocol for the detection of B.1.1.7 lineage of SARS-CoV-2 by using SYBR Green-Based RT-qPCR

**DOI:** 10.1101/2021.01.27.21250048

**Authors:** Fadi Abdel Sater, Mahmoud Younes, Hassan Nassar, Paul Nguewa, Kassem Hamze

## Abstract

**Background:** The new SARS-CoV-2 variant VUI (202012/01), identified recently in the United Kingdom (UK), exhibits a higher transmissibility rate compared to other variants, and a reproductive number 0.4 higher. In the UK, scientists were able to identify the increase of this new variant through the rise of false negative results for the spike (S) target using a three-target RT-PCR assay (TaqPath kit).

**Methods:** To control and study the current coronavirus pandemic, it is important to develop a rapid and low-cost molecular test to identify the aforementioned variant. In this work, we designed primer sets specific to SARS-CoV-2 variant VUI (202012/01) to be used by SYBR Green-based RT-PCR. These primers were specifically designed to confirm the deletion mutations Δ69/Δ70 in the spike and the Δ106/Δ107/Δ108 in the NSP6 gene. We studied 20 samples from positive patients, 16 samples displayed an S-negative profile (negative for S target and positive for N and ORF1ab targets) and four samples with S, N and ORF1ab positive profile.

**Results:** Our results emphasized that all S-negative samples harbored the mutations Δ69/Δ70 and Δ106/Δ107/Δ108. This protocol could be used as a second test to confirm the diagnosis in patients who were already positive to COVID-19 but showed false negative results for S-gene.

**Conclusions:** This technique may allow to identify patients carrying the VUI (202012/01) variant or a closely related variant, in case of shortage in sequencing.

## Introduction

As the pandemic of SARS-CoV-2 continues to affect the planet, researchers around the world are monitoring the virus and detecting acquired mutations that may lead to higher threats of spreading COVID-19 [1]. A new variant was recently reported in the UK as a Variant Under Investigation (VUI - 202012/01), belonging to the B.1.1.7 lineage [2]. Its rate of transmission is estimated to be >70%, and its reproductive number (Ro) seems to be up to 0.4 higher [3]. This variant harbors 14 non-synonymous mutations, 6 synonymous mutations and 3 deletions [2, 3]. A wide variety of diagnostic tests have been used by high-throughput national testing systems around the world, to monitor the SARS-CoV-2 infection [4]. The arising prevalence of new SARS-CoV-2 variants such as B.1.1.7 has become of great concern, as most of the existing RT-PCR tests will not be able to specifically distinguish these new variants because they were not designed for such a purpose. Therefore, public health officials most rely on their current testing systems and their sequencing results to draw conclusions on the prevalence of new variants in their territories [2, 5]. An example of such cases has been seen in the UK. In fact, scientists were able to identify the augmentation of the B.1.1.7 SARS-CoV-2 variant infection in the population through an increase in the S-gene target failure in their three target gene assay (N+, ORF1ab+, S-) when using the Applied Biosystems TaqPath RT-PCR COVID-19 kit (Thermo Fisher Scientific, Waltham, USA) that included the ORF1ab, S, and N gene targets [1, 6, 7].

In 24 December 2020, Thermo Fisher Scientific confirmed that the S deletion Δ69/Δ70 was in the area targeted by the TaqPath Kit. Whilst other variants with Δ69/70 are also circulating worldwide, the absence of detection of the S gene target increasingly appears to be a highly specific marker for the new variant [2, 7]. In addition, the European CDC recommended that multi-target RT-PCR assays that included an S gene target affected by the deletions could be used as a signal for the presence of the Δ69/Δ70 mutation for further investigations and could be helpful to keep tracking these mutant strains [8].

Genome sequencing is the gold method to confirm the new variant, but observational studies provide also stronger evidence if similar models are observed in multiple countries, especially when randomized studies are not possible. In Lebanon, surveillance data from Beirut Medical Center and Bahman Hospital showed a rapid and dramatic increase in S-negative profile in PCR testing for SARS-CoV-2 in the first twelve days in January, reaching approximately 60% of positive cases [9].

In 15 January 2021, in GISAID we found 374,613 SARS-CoV-2 sequences, among them 17,473 sequences belonged to the new variant (VUI 202012/01). It is important to notice that the co-occurrence of mutations Δ69/Δ70 in the spike and Δ106/Δ107/Δ108 in the NSP6, was present in this new variant. We then decided to use these two deletions as targets for our rapid and low-cost protocol, performing SYBR Green-Based RT-PCR. In this work, we propose primer sets that can be applied as a second step to confirm the diagnosis in cases that were already detected as positive to SARS-CoV-2.

It is well known that confirmatory diagnosis based on specific diagnostic biomarkers remains a great challenge allowing the further control and eradication of infections including COVID-19 [10]. Therefore, this novel method may lead to specifically identify individuals carrying the Δ69/Δ70 and the Δ106/Δ107/Δ108 mutations.

## Material and Methods

### Clinical specimens

20 clinical samples for SARS-CoV-2 positive patients, with Ct (cycle threshold) value < 30, were selected for this study from December 9 to January 10. All samples were previously tested positive by the Applied Biosystems™ TaqPath™ COVID-19 assay which targeted the RdRP, N and Spike genes in the Molecular Laboratories at Bahman Hospital and Beirut Cardiac Institute in the city of Beirut. 16 of these samples were S-negative and 4 were S-positive.

All patients provided written and signed informed consents

### RNA extraction and cDNA obtention

RNA was extracted from the clinical samples using Qiamp viral RNA mini kit (Qiagen). Total RNA was converted to cDNA using iScript cDNA synthesis kit (BioRad), following the manufacturer’s recommended procedures.

### Primer design

Primers were designed based on the recently available full sequence of the new variant VUI (202012/01) from the NCBI Reference Sequence Database (https://www.ncbi.nlm.nih.gov/nuccore/NC_045512). We utilized NCBI-Primer BLAST (https://www.ncbi.nlm.nih.gov/tools/primer-blast/) to design specific primers (Figure x). The primer sets were synthesized and delivered by Macrogen (Republic of Korea), they are listed in table 1.

**Table 1.**
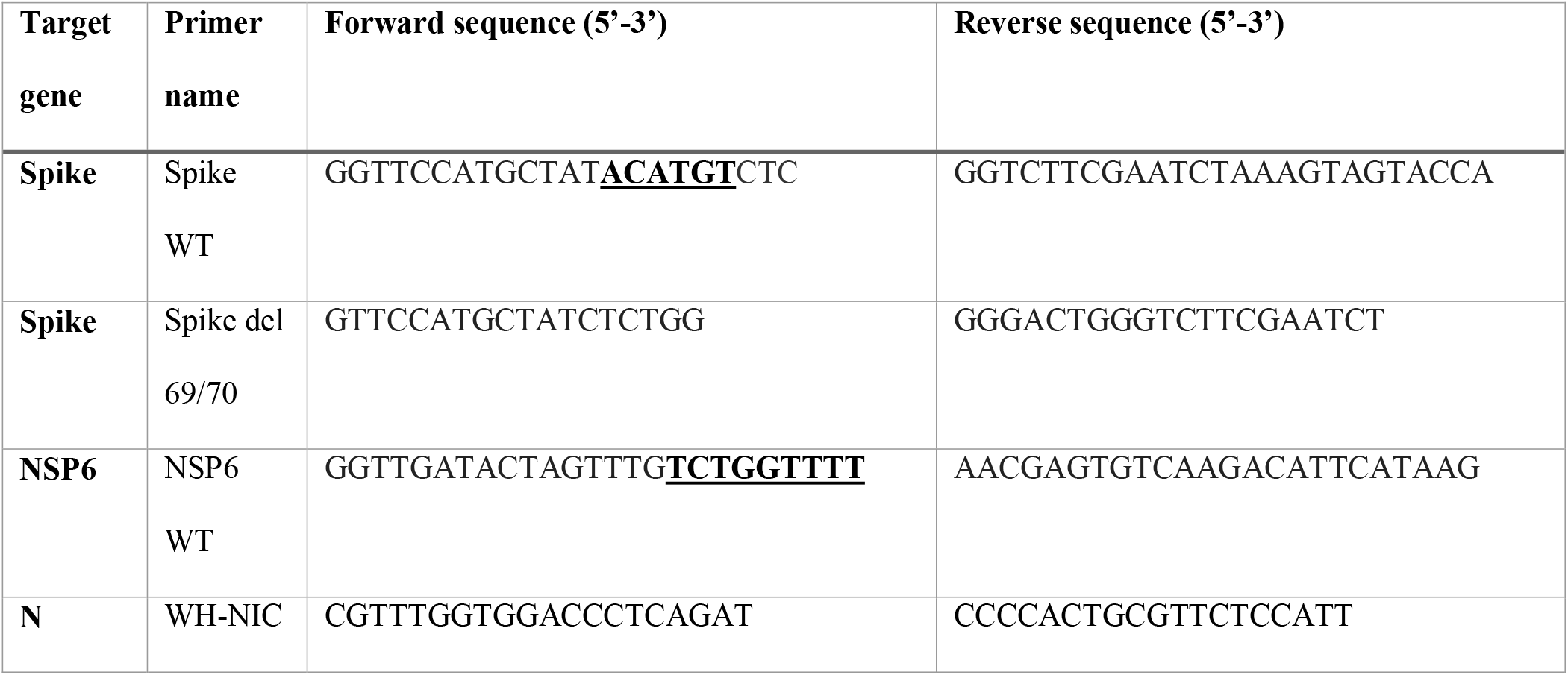
List of primer sets. Deleted nucleotides are in bold and underlined.

### SARS-CoV-2 rRT-PCR

Quantitative RT-PCR was carried out using iTaq universal SYBR green super mix (Bio Rad). In brief, each reaction consisted of a total volume of 20 μl containing 2 μL of each primer [10 pM/μL], 5 μl of cDNA, 10 μl SYBR Green super mix and 1 μL of Rnase free Water. Real-time PCR was performed using Bio Rad CFX96 Real□Time PCR Machine. The thermal cycling conditions used were as follows: 94 °C for 2 min, followed by 40 cycles of amplification at 94°C for 10 seconds, and 60°C for 1 minute. The reaction was completed by determining the dissociation curve of all amplicons.

## Results

In order to validate our method, we re-tested 20 samples that had been tested as positive for SARS-CoV-2 with Ct < 30, by the TaqPath kit. 16 samples with S-negative profile and four samples with S-positive profile (Table 2). N primer pairs were used as positive control, Spike WT and NSP6 WT primer pairs were used to detect the variants not harboring the deletion mutations Δ69/Δ70 in the spike nor the Δ106/Δ107/Δ108 in the NSP6, respectively. Spike del 69/70 primer pairs were designed to detect the deletions Δ69/Δ70 in the spike protein (Figure 1).

**Table 2.** The Ct values from the TaqPath Kit and SYBER Green-Based assay. Ct: cycle threshold; ND: not detected.

|  |  | Probe Based (TaqPath Kit) |  |  | SYBER Green- Based |  |  |  |
| --- | --- | --- | --- | --- | --- | --- | --- | --- |
|  |  | Ct values |  |  | Ct values |  |  |  |
|  | Samples | ORF1 | N | Spike<br>(S) | N<br>(positive<br>control) | NSP6<br>(ORF1) | Spike<br>(S) | spike del<br>69/70 |
| S-negative samples | 1 | 15 | 16 | ND | 15 | ND | ND | 16 |
|  | 2 | 25 | 25 | ND | 27 | ND | ND | 28 |
|  | 3 | 17 | 17 | ND | 19 | ND | ND | 20 |
|  | 4 | 15 | 16 | ND | 25 | ND | ND | 23 |
|  | 5 | 15 | 17 | ND | 21 | ND | ND | 22 |
|  | 6 | 23 | 22 | ND | 23 | ND | ND | 24 |
|  | 7 | 15 | 16 | ND | 21 | ND | ND | 21 |
|  | 8 | 22 | 23 | ND | 18 | ND | ND | 17 |
|  | 9 | 20 | 19 | ND | 26 | ND | ND | 26 |
|  | 10 | 19 | 21 | ND | 22 | ND | ND | 24 |
|  | 11 | 13 | 14 | ND | 18 | ND | ND | 19 |
|  | 12 | 14 | 14 | ND | 17 | ND | ND | 20 |
|  | 13 | 32 | 31 | ND | 33 | ND | ND | 31 |
|  | 14 | 20 | 21 | ND | 19 | ND | ND | 22 |
|  | 15 | 20 | 22 | ND | 25 | ND | ND | 27 |
|  | 16 | 17 | 17 | ND | 19 | ND | ND | 18 |
| S-positive samples | 17 | 24 | 24 | 24 | 25 | 26 | 27 | ND |
|  | 18 | 19 | 19 | 18 | 21 | 19 | 19 | ND |
|  | 19 | 23 | 23 | 25 | 26 | 27 | 28 | ND |
|  | 20 | 20 | 20 | 21 | 19 | 21 | 22 | ND |

**Figure 1.**
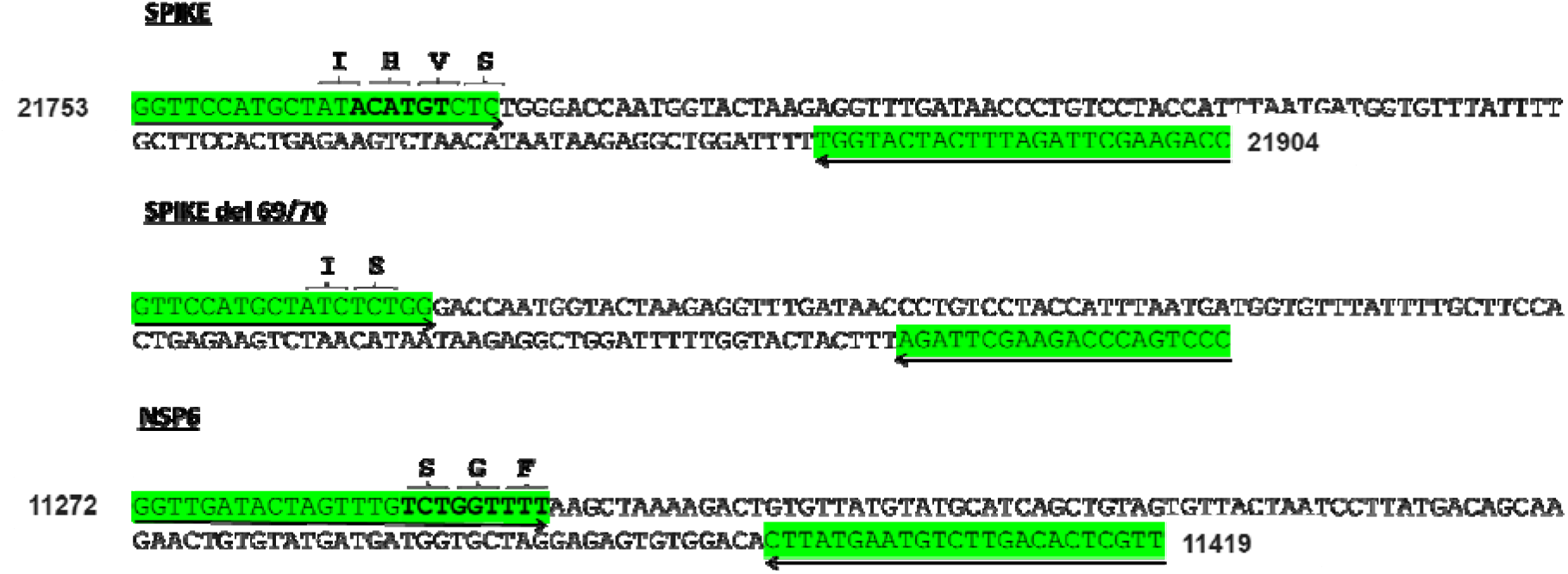
The Localization of the selected primers. Deleted nucleotides are written in bold.

Our results showed that the N-gene amplicons were detected in both, S-negative and S-positive profiles proving the role of this primer as a positive control (Table 2 and Figure 2), whereas the spike del 69/70-gene amplicons were detected only in the S-negative profile confirming the absence of the amino acids 69 and 70 in the spike protein of these samples (Figure 2. B, Figure 2), For the spike WT and NSP6 WT primers, amplicons of both, were detected only in the S-positive profile confirming the presence of the two amino acids 69 and 70 in the spike protein and the three amino acids 106, 107 and 108 in the NSP6 protein of S-positive samples (Figure 2.A). The Ct values of both TaqPath kit and SYBR Green-based RT-PCR were similar to a difference in Ct of 2.1 on average when comparing the Ct values of N gene in Probe-based and SYBR Green –Based PCR (Table 2).

**Figure 2.**
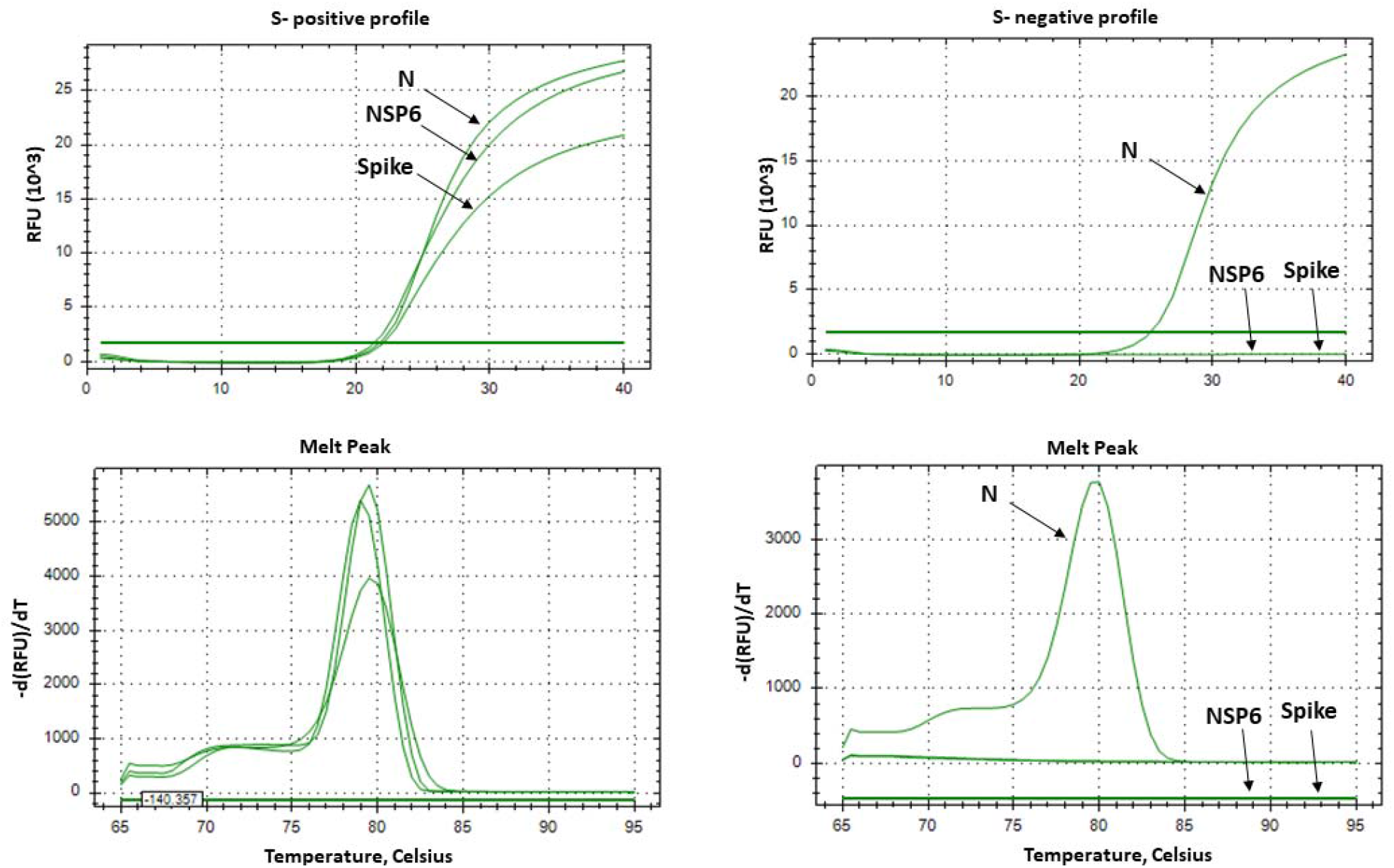
Real time PCR results, from SYBR Green-Based assay, with primers targeting the non-mutated N, Spike and NSP6 genes in S-positive and S-negative samples (N-gene is used as a positive control). Panel A show the amplification curves of the targeted regions in N, S and NSP6 genes in the S-positive samples. Panel B show the amplification curves of the targeted region in N gene in the S-negative samples. Panel C and D show the melting curves of the targeted regions in S-positive and S-negative samples, respectively. In panel D only one melting peak in S-negative samples, corresponding to the N gene.

The results of spike del 69/70 primers were fully concordant with spike WT and NSP6 WT primers, 100% of the S-negative profile had the Spike deletions Δ69/Δ70 (Figure 3. A), while 100% of the S-positive profile did not contain the deletions Spike Δ69/Δ70 and NSP6 Δ106/Δ107/Δ108 (Figure 2. A). Moreover, we confirmed the presence of only one PCR product by a unique melting peak for each primer pairs (Figure 2. C, D).

**Figure 3.**
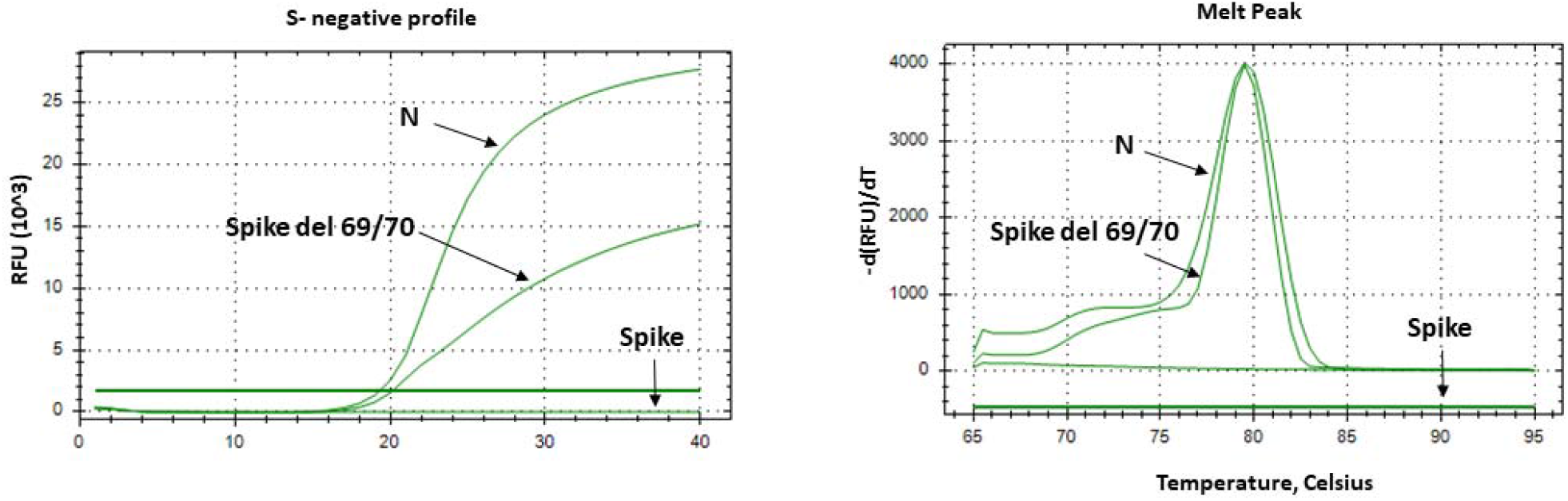
SYBR Green-Based PCR with primers targeting the mutated Spike del 69/70 genes in S-negative samples (N-gene is used as a positive control). Panel A show the amplification curves of the targeted regions in N, and Spike genes in the S-positive samples using three primer sets corresponding to N, Spike WT and Spike del 69/70. Panel B show the melting curves for the products amplified in S-negative samples by N and Spike del 69/70 primers only.

## Conclusion

Sequencing is the current gold standard diagnostic method to confirm the presence of any new variant, its efficient but a time consuming method and not accessible to all laboratories especially in developing countries. In this study, we have developed a rapid, low-cost, large-scale screening protocol for the detection of the deletions Δ69/Δ70 and Δ106/Δ107/Δ108. We hope that our efforts will be helpful and can contribute to the early detection of the new variant (VUI 202012/01), for the prevention of transmission and early intervention. This protocol should not be limited to this variant, but also for any other future variant to come, just by designing the appropriate primers.

## Data Availability

All data are present in the manuscript

## ACKNOWLEDGEMENTS

This work was supported by the Lebanese University and Fundación La Caixa (LCF/PR/PR13/11080005), Fundación Caja Navarra, Fundación Roviralta, Ubesol, Inversiones Garcilaso de la Vega, COST Actions CA18217 and CA18218, and EU Project uncover (Grant/Award Number: 101016216).

